# Distinct patterns of cognitive outcome in young children with autism spectrum disorder receiving the Early Start Denver Model

**DOI:** 10.1101/2021.04.05.21254908

**Authors:** Godel Michel, Robain François, Kojovic Nada, Franchini Martina, Wood de Wilde Hilary, Schaer Marie

## Abstract

Evidence-based, early intervention significantly improves developmental outcome in young children with autism. Nonetheless, there is high interindividual heterogeneity in developmental trajectories during the therapy. It is established that starting intervention as early as possible results in better developmental outcomes. But except for younger age at start, there is no clear consensus about behavioral characteristics that could provide a reliable individual prediction of a child’s developmental outcome after receiving an early intervention. In this study, we analyze developmental trajectories of preschoolers with autism who received 2 years of intervention using the Early Start Denver Model (ESDM) approach in Geneva, Switzerland in an individual setting (n = 55, aged 28.7 ± 5.1 months with a range of 15 – 42). Our aim was to identify early predictors of response to treatment. We applied a cluster analysis to distinguish between 3 groups based on their cognitive level at intake, and rates of cognitive change over the course of treatment. The first group of children only had a mild cognitive delay at intake and nearly no cognitive delay by the end of treatment (Higher Cognitive at baseline: HC). The children in the two other groups all presented with severe cognitive delay at baseline. However, they had two very different patterns of response to treatment. The majority significantly improved developmental scores over the course of treatment (Optimal Responders: OptR) whereas a minority of children showed little to no improvement (Minimal Responders: MinR). Further analyses showed that children who ended up having an optimal two-year treatment outcome (OptR) were characterized by higher adaptive functioning at baseline combined with rapid developmental improvement during the first 6 months of intervention. Inversely, less significant progress by the sixth month of intervention was associated with a less optimal response to treatment (MinR).

## Introduction

Autism spectrum disorder (ASD) is characterized by impairments in communication and social interactions, along with restricted and repetitive behaviors (American Psychiatric Association, 2013). Over the last three decades, several comprehensive, evidence-based early intervention (EI) approaches have been developed for young children with ASD, with the aim to improve their social communication, cognitive functioning, and adaptive skills (Dawson et al., 2010; Freitag et al., 2012; Koegel et al., 1999; Lovaas et al., 1974; Prizant et al., 2006; Schreibman et al., 2015). The principles of EI usually comprise a significant number of hours (usually more than 15 hours per week) as well as an early age of onset (usually younger than 4 or 5 years old) (Corsello, 2005). Systematic reviews and meta-analyses showed positive effects of EI on cognition, adaptive skills and communication at the group level (Eldevik et al., 2009; Fuller & Kaiser, 2020). Nevertheless, many EI studies have reported a relatively heterogeneous response to these interventions, where most children show significant improvements, while others make smaller gains (Howlin et al., 2009; Warren et al., 2011). Despite important efforts to better understand variables affecting treatment response, it is currently not possible to predict to what extent a child will respond to intervention based on his or her behavioral characteristics at intake (Vivanti et al., 2014). In the current therapeutic context and in the absence of additional knowledge about individual predictors of outcome, many authors suggest that intensive early intervention should be an intervention of choice for young children diagnosed with ASD (Fuentes et al., 2021; Zwaigenbaum et al., 2015) regardless of their specific behavioral or symptom profile. Yet, in the global framework of precision medicine (König et al., 2017), there is an urge to develop more individualized guidelines for intervention in ASD. Given the importance of providing effective programming for children with ASD as early as possible, and because of the costs and parental investment associated with early intervention, it is crucial that we move away from a “one size fits all” service provision model, and find ways to tailor a child’s intervention to their specific needs, choosing therapy approaches based on the child’s individual profile at diagnosis (Cidav et al., 2017; Penner et al., 2015; Peters-Scheffer et al., 2012).

During the last decade, Early Start Denver Model (ESDM) has emerged as a promising Naturalistic Developmental Behavioral Interventions (NDBI) (Schreibman et al., 2015). NDBIs represent a category within the broader context of EIs, as discussed by Vivanti et al (Vivanti & Stahmer, 2021). Briefly, NDBIs designates approaches that integrate the methods derived from behavioral learning and developmental science. Main principles include varying the stimuli for learning, using the activities the child enjoys the most and emphasis put on developmental prerequisites. Within NDBIs, ESDM is notably illustrated by its overall effectiveness, its emphasis on natural environment teaching, comprehensive learning objectives and parental involvement. ESDM intervention has originally been implemented in an individualized setting (one therapist for one child, I-ESDM), but other applications have been developed such as G-ESDM where one therapist works with a little group of children and P-ESDM where parents/caregivers actually provide the intervention under supervision. In their 2010 landmark randomized controlled trial (RCT) (Dawson et al., 2010), Rogers and Dawson reported a mean increase of 18 IQ points in a sample of 24 toddlers with ASD receiving the I-ESDM intervention over two years. Numerous studies have replicated these results (for a review see (Fuller & Kaiser, 2020)), highlighted a good reproducibility in different contexts such as the European one (Colombi et al., 2018; Department for Child and Adolescent psychiatry, Centre Hospitalier le Vinatier, Bron, France et al., 2019; Devescovi et al., 2021) and demonstrated the cost-effectiveness of ESDM intervention (Cidav et al., 2017; Penner et al., 2015). Overall, the ESDM approach has been shown to significantly increase cognitive, communication and adaptive skills at the group level (Fuller et al., 2020). However, the inter-individual variability in child response to treatment (RTT) is high, as with all types of EI (Contaldo et al., 2020). To date, research about RTT in ESDM remains sparse and most studies focusing on homogeneous and individualized therapy settings comprised limited sample size. Younger age at start has emerged as an important moderator of optimal outcome, probably due to higher brain plasticity (Lombardo et al., 2021; Webb et al., 2014). Age left aside, there are no behavioral child that are recommended by any international guidelines as a reliable individual predictor of RTT in ESDM, despite many attempts to identify some (Fuentes et al., 2021; Lord et al., 2020). E.g., Vivanti et al. (Vivanti et al., 2013) attempted to identify predictors of RTT in children receiving G-ESDM intervention. Their study showed that developmental gains after one year of treatment were best predicted by higher imitation skills, goal understanding and more advanced skills in the functional use of objects at baseline. This study offered insight into how children with certain baseline competencies might progress faster in a G-ESDM setting. However, outcomes were assessed after only one year of intervention and baseline measures used in this study were based on original tasks (i.e., specially developed for this study and not available in the common practice), making its results poorly reproducible. In addition, its group setting makes its conclusion hardly generalizable to the canonical individualized setting of ESDM. Besides, some authors identified that lower cognitive level at baseline could be related to higher RTT, although this effect could be biased by a larger potential for gain in children with very low cognitive profile (Devescovi et al., 2016; Robain et al., 2020). This brief review shows that various behavioral characteristics (e.g. global cognitive level or imitation skills) at baseline modulate the outcome of an ESDM intervention. Nevertheless, none of these parameters has reached the status of being a reliable predictor of individual response to ESDM intervention recommended by international guidelines yet (Fuentes et al., 2021). It is therefore currently not possible to know whether a child will respond or not to the ESDM intervention when advising it. Yet, the identification of characteristics that promote the response to a specific intervention could in the future be of great help to the clinical practice when referring a child to one EI or another. Similarly, new approaches or goals could also be implemented to promote the emergence of these predictors to create cascading effects on children’s intervention response. Great interindividual heterogeneity in response to intervention has been identified as a major limitation to this quest (Vivanti et al., 2014). A promising way of dealing with this heterogeneity relies on moving from a whole-group approach to the identification of distinct subgroups exhibiting specific patterns of response to intervention (Ousley & Cermak, 2014).

In the present study, we aim to identify early children’s behavioral characteristics that could serve as predictors of outcome after receiving a specific and homogeneous NDBI (here, I-ESDM). To do so we explored the developmental trajectories of 55 preschoolers with ASD who completed two years of individualized and intensive (20 hours per week) ESDM intervention available in Geneva, Switzerland. We used a longitudinal single group design without a control population, similar to previous studies in the field (Contaldo et al., 2020; Lombardo et al., 2021; Vivanti et al., 2013). Indeed, because of ethical as well as logistic considerations, a random referencing to either the ESDM intervention program or any other community treatment was not achievable. We first investigated if our sample’s outcome data, in terms of cognition, symptom severity and adaptive functioning, reflected findings described in the ESDM literature. We then parsed the heterogeneity in our sample’s outcome by using cluster analysis (CA) and cognitive scores as the main outcome measure. CA highlighted three different groups based on cognitive outcome. We further explored baseline differences as well as early rates of change between the three groups to identify potential predictors of two-year treatment outcome.

## Method

### Participants

Our original sample included 61 participants who completed two years of ESDM intervention in Geneva, Switzerland. Six participants were not included in the analyses because of missing data regarding their developmental assessment at baseline and one participant because of missing data at the end of the intervention. Missing evaluations were all caused by logistical issues (e.g. evaluation material not available at this time) and not because of children characteristics (e.g. invalid evaluation because of the child’s behavior). Full description of the six excluded children is provided in Supplementary Table 3. There was no significant diference between the excluded participants and the final sample. Our analyses were thus based on the data collected from 55 participants (see Table 1).

**Table 1:** Sample characteristics over the two years of ESDM intervention. Scores at 6 months (in italic) are indicative and were not used in the repeated measure (R.M.) ANOVA. Scores with a significant difference are highlighted in bold; *\*p<.05 ** p<.01 ***p<.001*. G: *Greenhouse-Geisser correction applied, ESDM = Early Start Denver Model*

| MEASURE | At Baseline | + 6 months | + 12 months | +24 months | Pval (R.M. ANOVA) | Partial Eta Squared | 0 - 24mo | 0 - 12mo | 12mo - 24mo |
| --- | --- | --- | --- | --- | --- | --- | --- | --- | --- |
| **Clinical Description** |  |  |  |  |  |  |  |  |  |
| ADOS CSS total [Mean (SD)] | **8.0 (1.9)** |  | **6.6 (2.0)** | **6.6 (2.7)** | **<.001\*\*\*** | .169 | **0.002\*\*** | **<0.001\*\*\*** | 1.000 |
| ADOS CSS SA | **7.6 (2.0)** |  | **5.3 (1.7)** | **5.5 (2.5)** | **<.001\*\*\* (G)** | .304 | **<0.001\*\*\*** | **<0.001\*\*\*** | 1.000 |
| ADOS CSS RRB | **8.0 (2.0)** |  | **9.1 (1.3)** | **8.4 (2.7)** | **<.007\*\* (G)** | .094 | **0.975** | **<0.001\*\*\*** | 0.122 |
| ADI-R subdomains [Mean (SD)] (n = 51) |  |  |  |  |  |  |  |  |  |
| ADI-R Social interactions | 14.2 (5.2) |  |  |  |  |  |  |  |  |
| ADI-R RRB | 4.0 (2.2) |  |  |  |  |  |  |  |  |
| VABS-II Adaptative Behavior Composite [Mean (SD)] (n=48) | 79.9 (9.4) | *81.3 (12.0)* | 80.7 (12.3) | 83.3 (16.1) | **0.046\* (G)** | **0.073** | **0.152** | **1.000** | **0.044\*** |
| VABS-II socialization | 80.6 (9.9) | *81.0 (9.9)* | 80.7 (10.2) | 80.4 (12.6) | 0.967 (G) |  |  |  |  |
| VABS-II communication | **75.6 (12.3)** | *80.5 (14.8)* | **84.0 (17.7)** | **87.6 (19.0)** | **<0.001\*\*\* (G)** | .377 | **<0.001\*\*\*** | **<0.001\*\*\*** | **0.008\*\*** |
| VABS-II daily living skills | 83.57 (12.4) | *85.0 (11.6)* | 84.8 (11.8) | 84.5 (16.8) | 0.726 (G) |  |  |  |  |
| VABS-II motor skills | 89.5 (11.6) | *89.4 (11.0)* | 89.9 (10.6) | 90.1 (15.2) | **0.920 (G)** |  |  |  |  |
| Composite DQ [Mean (SD)] | **60.1 (17.6)** | *71.7 (20.9)* | **77.0 (25.0)** | **80.0 (28.1)** | **<0.001\*\*\*** | .325 | **<0.001\*\*\*** | **<0.001\*\*\*** | 1.000 |
| Fine Motricity DQ | **74.3 (17.0)** | *76.1 (16.0)* | **76.1 (18.4)** | **81.7 (25.5)** | **0.035 (G)\*** | .063 | **0.088** | **1.000** | **0.132** |
| Visual Reception DQ | **74.6 (22.9)** | *85.3 (23.5)* | **88.6 (30.8)** | **88.6 (30.7)** | **<0.001\*\*\* (G)** | .150 | **0.003** | **<0.001\*\*\*** | 1.000 |
| Expressive Language DQ | **44.0 (19.2)** | *56.8 (25.3)* | **66.4 (29.3)** | **68.7 (28.0)** | **<0.001\*\*\* (G)** | .447 | **<0.001\*\*\*** | **<0.001\*\*\*** | 0.782 |
| Receptive Language DQ | **47.4 (26.2)** | *68.8 (29.0)* | **76.8 (30.1)** | **79.0 (33.6)** | **<0.001\*\*\* (G)** | .432 | **<0.001\*\*\*** | **<0.001\*\*\*** | 1.000 |
| **Demographics** |  |  |  |  |  |  |  |  |  |
| Chronological age [months Mean (SD)] | 28.7 (5.1) |  |  |  |  |  |  |  |  |
| Gender [females Number ( percentage)] | 7 (12.7 %) |  |  |  |  |  |  |  |  |
| Parental Education [Number (percentage)] (n=46) |  |  |  |  |  |  |  |  |  |
| Elementary School or High School | 21 (38.2%) |  |  |  |  |  |  |  |  |
| University or PhD | 33 (60.0 %) |  |  |  |  |  |  |  |  |
| Household Income [Number (percentage)] (n=45) |  |  |  |  |  |  |  |  |  |
| <60k | 14 (25.5%) |  |  |  |  |  |  |  |  |
| 60-140k | 18 (32.7%) |  |  |  |  |  |  |  |  |
| >140k | 21 (38.2 %) |  |  |  |  |  |  |  |  |

There was no exclusion criteria based on co-occurring somatic, neurologic or genetic disorder, as long as they were not affecting the validity of behavioral measures (e.g., major cerebral palsy). There was no systematic genetic or neurological screening done in our protocol. Genetic, somatic and neurologic diagnosis were screened with parental questionnaires. To our knowledge, no children were affected by any neurologic condition (e.g., epilepsy) diagnosed by a neurologist following active consultation by parents. No parents reported any diagnosis of major somatic disorders that could have affected the validity of behavioral measures. Twenty-four participants’ parents reported having met a clinical geneticist. Four of them reported a genetic finding that could be “causative for ASD” according to the geneticist’s report.

All children were referred to the intervention program after receiving a clinical diagnosis of ASD according to Diagnostic and Statistical Manual of mental disorders, 5^th^ edition (American Psychiatric Association, 2013) criteria and Autism Diagnosis Observation Schedule-Generic (ADOS-G) (Lord et al., 2000) or 2^nd^ edition (ADOS-2) (Lord et al., 2012) diagnosis cut-offs. For children that were administered the ADOS-2 Toddler module (which does not provide a diagnostic cut-off) at baseline, the “mild to moderate concern for ASD” cut-off had to be overreached. All children assessed with a Toddler module at baseline met the diagnosis cut-off (using ADOS-2 module 1 or 2) on their visit one year later, even though this was not an inclusion criterion for our study.

Enrollment in the intervention program was also conditioned by an age criterion: participants had to be able to participate for two full years prior to age of school entry. In Geneva, a child has to be 4 years old by July 31^st^ to enter school in August of the same year. In our sample, one child was too old (42 months old at baseline) to meet this criterion but was still enrolled in the program as there was an available position. This results in a sample that is fairly homogeneous in age at start (28.7 ± 5.1 months, see Main Table).

At least one parent had to be fluent in either French or English. Therapists fluent in both these languages were available to provide intervention, follow-up and parental coaching. The latest census in Geneva (Office Fédéral de la statistique / Office cantonal de la statistique, 2021) reports that 92.3% of the population use either French or English as a first language. We must add to this percentage the people fluent in French or English as a second language. Thus, the vast majority of the population in Geneva was eligible for the intervention program based on the language inclusion criterion. Besides, there has been increasing concerns about socio-economic representativeness of the samples used in EI research (Mirenda et al., 2022; Nahmias et al., 2019). To date the majority of ESDM studies are based on a white population with high parental income and a college educated background (S. J. Rogers et al., 2020). Geneva has a very culturally diverse population and the costs of the ESDM intervention program are almost completely covered) (Office Cantonal de la statistique (OCSTAT), 2017). As a result, our sample is fairly representative of Geneva’s residents socio-economic characteristics thus providing results with a very high degree of cultural and socio-economic generalizability compared to most studies in the field (see Main Table).

Ultimately, enrollment also depended on place availability at time of referral. The parents of all participants gave their written informed consent to the research protocol that was approved by the institutional review board of the University of Geneva. All participants were assessed in the context of the ongoing longitudinal Geneva Autism Cohort study. Twenty-two children from this same sample were already included in a previous study measuring outcome after one year of ESDM intervention (Franchini et al., 2018). Baseline evaluations were completed at the start of the intervention and comprised behavioral measures that are detailed below. Parents also filled out questionnaires regarding medical history, as well as demographic information detailed below. Children were then assessed at 3 other time points at 6, 12 and 24 months of therapy, for a total of 4 assessments. Post-intervention data about subsequent school placement and support needs were collected. Children went onto either regular educational classrooms with varied levels of in-class paraprofessional support or special education classrooms.

### Intervention

The 55 participants were enrolled in one of the 4 units of the *Centre d’Intervention Précoce en Autisme* (CIPA) in Geneva, Switzerland (Fondation Pôle Autisme (http://www.pole-autisme.ch) & Office Médico-Pédagogique), where they received 20 hours a week of daily, individual intervention sessions using the Early Start Denver Model (ESDM). The ESDM is a comprehensive, evidence-based early intervention approach that promotes child learning through naturalistic developmental, and behavioral techniques (SJ. Rogers & Dawson, 2010; Schreibman et al., 2015). Parents of the participants were provided with 12 hours of once-a-week parent coaching sessions in the use of the ESDM model at the start of their child’s program, and continued parent support sessions as needed throughout the two-year period. The children were evaluated every 3 months using the Early Start Denver Model Curriculum Checklist for Young Children with Autism (ESDM-CC) to establish targeted and measurable learning objectives. The intervention services were provided by graduate-level therapists (at least Master’s degree), who were trained within the CIPA program in the use of the ESDM approach, meeting ESDM fidelity on the ESDM Fidelity Rating System (SJ. Rogers & Dawson, 2010). Today, the team consists of 20 credentialed ESDM therapists, and the program is overseen by an ESDM certified trainer. Importantly, university background, ESDM training, fidelity rating assessment and supervision by certified trainer does not differ across the four units. The separation in four units is essentially administrative and therapists are all part of the same team sharing the same supervisors, applying identical practice.

### Measures

The ADOS (which refers to the ADOS-G and its later version, the ADOS-2), is a standardized assessment which comprises a series of semi-structured social presses aimed to elicit and measure ASD symptoms (Lord et al., 2000, 2012). The schedule comprises 5 different modules, adapted to the person’s age and level of language. The calibrated severity score (CSS) was used to compare the total severity score as well as the restricted and repetitive behaviors (RRB) and social affect (SA) symptoms severity scores (Gotham et al., 2009; Hus et al., 2014). The ADOS were administered by a trained examiner and filmed. The members of the team who rated the video recordings were not implicated in the delivery of the ESDM intervention.

The Mullen Scales of Early Learning (MSEL) is a standardized assessment for children aged from birth to 68 months (Mullen, 1995). It measures the child’s development in five developmental domains: expressive language (EL), receptive language (RL), visual reception (VR), fine motor (FM), as well as gross motor skills (GM).

The Psychoeducational Profile – third edition (PEP-3) is a standardized assessment tool that evaluates cognitive, motor, and adaptive domains in children 2 to 7 years of age (Schopler et al., 2005). These domains include EL, RL, FM and cognitive verbal and preverbal (CVP). The PEP-3 as well as the MSEL were administered by psychologists following the standard instructions of both evaluations.

Developmental quotient scores (DQ) were computed for each subdomain of the MSEL by dividing the individual developmental age by the chronological age and multiplying by 100 as described in 2006 by Lord and colleagues (Lord et al., 2006). The composite DQ was computed by calculating the average of all four subdomains’ developmental ages, then dividing by the chronological age and multiplying by 100. Similarly, DQ scores were computed for the subdomains of the PEP-3 that assess domains equivalent to those of the MSEL, namely EL, RL, CVP and FM. The PEP-3 composite DQ was derived using the same formula as described for the MSEL, and has already been used for the PEP-3 subdomains in previous studies (Franchini et al., 2018). For our analyses of cognitive skills, we used the MSEL Early Learning Composite DQ. Since the MSEL was not administered for some participants (n = 7 at baseline, n = 7 after 6 months of therapy, n = 3 after 12 months of therapy, n = 2 after 24 months of therapy) we replaced the missing DQ scores by their equivalent DQ scores derived from the PEP-3. It is important to keep in mind that DQ is normalized for the age at the time of evaluation. Hence, a DQ that remains stable over time does not reflect stagnation but rather continued developmental progress. Also, a loss of DQ over time does not necessarily imply regression (a loss of skills) but rather slower skill acquisition, leading to a widening of the gap between the child’s current abilities and what would be expected in typical development.

The Vineland Adaptive Behavior Scales – 2^nd^ edition (VABS-II) is a semi-structured interview administered by a trained clinician that assesses a person’s adaptive behavior (Sparrow et al., 2005). The domains assessed comprise communication, socialization, daily living skills (DLS) and motor skills. An overall adaptive behavior composite score (ABC) of all these 4 domains is computed.

The ADOS, VABS-II, PEP-3 and MSEL were administered at baseline, after 12 months and after 24 months of therapy. Assessment at six months only comprised the VABS-II and the MSEL.

We measured participant socio-economic using the total household yearly income and the highest level of education achieved by parents at baseline. The household income was divided into three subgroups that are detailed in Table 1. Parental educational level was first coded using the seven categories of the four-factor index of social status developed by Hollingshead (Hollingshead, 1975). We then divided these categories into two groups: 1) elementary school or high school completed, and 2) college and/or graduate degree completed.

### Rate of change

For all behavioral measures acquired longitudinally (ADOS, VABS-II and DQ), we computed an individual rate of change using the following Symmetrized Percent Change (SPC) formula:

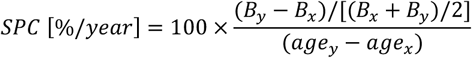

Where *B*_*x*_ and *B*_*y*_ represent the behavioral measure acquired when the participant was aged of *age*_*x*_ and *age*_*y*_ respectively. In other words, SPC is the behavioral difference between two timepoints relatively to the mean of the scores across these two timepoints, then divided by the time interval (in years). This results in a yearly rate of change that can be expressed as a percentage when multiplied by 100. The main advantages of using symmetrized measures of change over absolute differences (such as *B*_*y*_ − *B*_*x*_) or non-symmetrized percentages (such as (*B*_*y*_ − *B*_*x*_) / *B*_*x*_) comprise increased statistical robustness, higher reliability in small samples, limited sensitivity to outliers and equivalent consideration of both *B*_*x*_ and *B*_*y*_ measures (Berry & Ayers, 2006). Also, SPC was chosen over absolute difference because it is scaled for the global developmental level of the child. More specifically, the clustering algorithm described below would consider an absolute difference of 10 DQ points in a child with an initial DQ of 90 as equivalent to a gain of 10 points in a child with a 60 composite DQ at baseline. In children with ASD and low DQ, small absolute gains have a larger impact in their adaptive behavior compared to their peers with higher DQ (Green & Carter, 2014). Hence, measuring rates of change relatively to each participant global developmental level as SPC does appears more clinically meaningful.

### Statistical Analyses

IBM®SPSS® Statistics v26.0.0.0 for macOs (Armonk, NY: IBM Corp.) was used for all analyses. Statistical significance threshold was set at alpha = 0.05. Graphs were obtained with Prism® v8.3.0 (GraphPad Software, La Jolla California USA, www.graphpad.com) and Matlab R2018b for MacOs (MathWorks).

To test for an effect of time a repeated measure ANOVA was performed on the whole sample for each longitudinal behavioral measure using the scores collected at baseline, 12 months, and 24 months after the start of the intervention services. Greenhouse-Geisser correction was applied whenever the assumption of sphericity was violated according to Mauchly test.

Until now, methodological strategies to identify intervention-specific predictors of EI outcome include whole-sample correlations between baseline and outcome measures (Klintwall & Eikeseth, 2012; Vivanti et al., 2013), comparison between subgroups defined based on an arbitrary cut-off such as rapid vs slow learners (Sallows & Graupner, 2005) or best vs non-best outcome (Perry et al., 2008; Sallows & Graupner, 1999). A promising alternative relies on the identification of distinct phenotypic subgroups within ASD (Ousley & Cermak, 2014). Defining more homogeneous subgroups based on behavioral characteristics in a data-driven manner can be achieved by applying cluster analyses (CA), a strategy that has already been used in ASD preschool studies (for a review see (Ousley & Cermak, 2014). To date, CA has only been applied once on children with ASD participating in an EI program (Applied Behavioral Analysis: ABA) with a special focus on language development (Frazier et al., 2021). We here performed a cluster analysis (CA) using cognition (assessed with the composite DQ measure) as our main outcome measure. There are several reasons why we chose DQ over other parameters. First, it is generally the main outcome measure reported in early intervention studies, and it displays the most variability (Dawson et al., 2010; Landa, 2018; Smith et al., 2000). Second, cognition has been shown to be the domain that improves the most after early intervention (Rodgers et al., 2021). Third, studies investigating possible ASD subtypes within ASD have shown that the most salient group differences emerge when categorized by cognitive skills (Witwer & Lecavalier, 2008). We used a *k*-means clustering approach to identify subgroups in terms of DQ trajectories with a maximal number of iterations set to 10 (Hartigan & Wong, 1979). We chose two variables that capture individual DQ trajectories: the composite DQ at baseline and the composite DQ SPC over the two-year intervention period. To objectively determine the number of clusters *k* we used a two-step clustering approach as suggested by Kodinariya (Kodinariya & Makwana, 2013). We used the two-step clustering algorithm developed by Chiu et al. (Chiu et al., 2001) as it is implemented in IBM®SPSS® Statistics. Briefly, this method firstly divides the sample into a set of sub-clusters through a sequential approach and secondly merges the sub-clusters through a hierarchical technique based on the log-likelihood distance between them. Finally, the Akaike’s information criterion is used to objectively determine the optimal number of clusters.

The cluster analysis (CA) yielded 3 optimal clusters based on the baseline composite DQ and the composite DQ SPC over 2 years (Fig 1) (with silhouette measure of cohesion and separation equal to 0.6). The ANOVA revealed that one of these clusters exhibited significantly greater composite DQ at baseline compared to the others and was therefore named “higher cognitive at baseline” (HC, n = 20). Its average DQ at baseline was 78.6 ± 10.9 with a range between 64.4 and 107.9 with a SPC of 9.9 ± 5.8 %/yr. This corresponds to an average 18.3 gain for a final DQ of 96.9 ± 14.3 with a range between 64.2 and 124.3. The second “optimal responders” cluster (OptR, n = 24) was characterized by high rates of progress within the two-year program. DQ at baseline was 51.5 ± 10.7 with a range between 21.9 and 66.8, and its average SPC was 23.8 ± 7.9 %/year. This corresponds to an average 34.6 gain for a final DQ of 86.2 ± 20.8 with a range between 32.5 and 130.3. The third “minimal responders” cluster (MinR, n = 11) was characterized by decreased rates of progress compared to the two other clusters with an average SPC of -11.5 ± 12.0 %/yr. Its composite DQ at baseline was 44.9 ± 8.1 with a range between 31.7 and 59.1. The average loss was 9.1 for a final DQ of 35.8 ± 8.9 with a range between 24.5 and 58.0. The OptR and MinR subgroups did not differ in composite DQ at baseline, with an average of 51.5 and 44.9 respectively. Together, they form a group of children with lower cognitive scores (LC) at baseline. Cluster differences over composite DQ at baseline and composite DQ SPC are illustrated on Fig 1. Detailed analyses are reported in supplementary Table 1 and 2.

**Figure 1:**
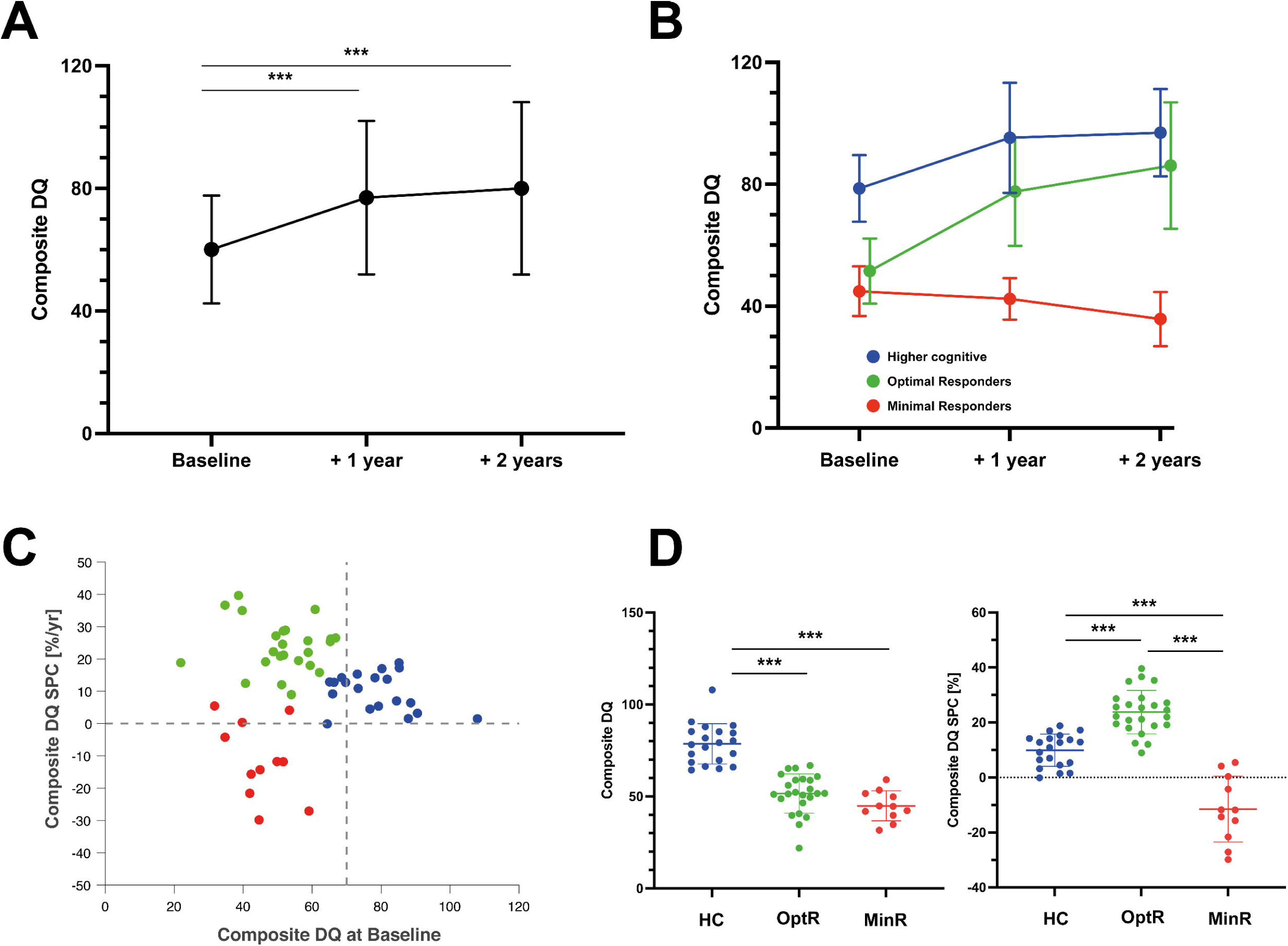
**A**. Composite DQ trajectory of the total sample over the two years of intervention. Significant results of repeated measure ANOVA are displayed. **B**. Composite DQ trajectories of the three subgroups parsed by the cluster analysis. **C**. Individual values of the two measures used in the clustering analysis algorithm (composite DQ at baseline and DQ SPC over the two years of interventions). Color code represents the cluster membership of each participant after the application of the cluster analysis. **D**. Differences between the three subgroups on the two measures that were used to parse them. \*\*\**p*<.001. DQ: Developmental Quotient. HC: Higher Cognitive. MinR: Minimal Responders. OptR: Optimal Responders. SPC: Symmetrized Percent Change.

Demographic, socio-economic measures and behavioral measures at baseline were compared between clusters using one-way analysis of variance (ANOVA) or chi-square test. We used a Bonferroni correction for multiple testing on the subdomains of the same clinical evaluation (e.g., the subdomains of the VABS-II), setting the statistical significance at 0.05/number of subdomains. When an ANOVA reached statistical significance, post-hoc comparisons between clusters were performed using multiple T-tests with Bonferroni correction and statistical significance set at 0.05/number of clusters.

We then applied the same strategy to compare the SPC between clusters. We performed analyses on the following SPC: from baseline to 6 months, from baseline to 12 months, from baseline to 24 months of therapy.

Finally, we focused on the two LC clusters which showed no differences in their composite DQ at baseline to explore whether any other behavioral measure could help classifying them. To do so, we used binary logistic regression models. More specifically, we selected all behavioral measures that differed between OptR and MinR on post-hoc T-tests at baseline. Then, we performed a multivariate logistic regression using the selected measures. Whenever a composite score and one or more subdomain scores of the same test were selected, we preferred the composite measure to minimize potential collinearity between variables in the model. Then, we used the same strategy for the SPC measures during the 6 first months of intervention, and ultimately with those of the 12 first months.

### Sample Size

Once the three clusters solution obtained, we were able to compute the estimated power to detect differences between groups. Based on a sample of 55 children divided in three clusters and assuming an alpha of 0.05 using ANOVA, we calculated 80% power to detect group differences of at least 0.430 effect size.

## Results

### Whole sample trajectories

Descriptive measures collected at each visit are reported in Table 1 for the total sample. The children were aged from 15.3 to 42.0 months at the beginning of the intervention (average:

28.7 ± 5.1 months). The average composite DQ of the entire group at baseline was 60.1 ± 17.6 (range: 21.9 – 107.9). As a group, all 55 children receiving ESDM showed a significant decrease in their total level of symptom severity (ADOS CSS) (see Table 1). This improvement was driven by a decrease in the Social Affect (SA) domain. On the contrary, the RRB symptom severity increased over time. We found that these changes occurred mainly during the first year of intervention and that CSS (both RRB and SA) were stable during the second year of intervention. In parallel, participants’ developmental scores improved. This improvement was significant in all subdomains (i.e., FM, VR, RL and EL). As for the measures of symptom severity, all changes in cognition were significant during the first year of therapy but not the second one, except for FM rates of change during first year that did not reach significance level in post-hoc analyses. Finally, increase in DQ was accompanied by an improvement of adaptive functioning as measured by the VABS ABC. This increase was significant during the second year of intervention only. More precisely, participants made significant gains in the communication subdomain which occurred both during the first and the second year of intervention. All statistically significant results are detailed in Table 1. Concerning the type of schooling after the intervention, 35 participants (63.6%) joined a public regular education classroom with individual educational support. One participant (1.8%) joined a regular education classroom without any support. Four children (7.3%) entered a private school that provided in-class support in a regular education classroom. Finally, 15 participants (27.3%) entered special education program within the public-school system.

### Parsing the heterogeneity in treatment response

#### Difference between the three subgroups at baseline

We found no differences between clusters for parental educational attainment or household income (see Supplementary Table 1). Inclusion procedure resulted in a sample that was homogeneous in age at baseline (28.7 ± 5.1 months). Yet we compared age at baseline between groups to exclude this variable as a confound factor and found no difference regarding age at baseline. When looking at DQ at baseline, we found that HC showed higher scores in all DQ subdomains compared to both other clusters. Considering adaptive behavior, HC exhibited higher scores in ABC as well as in the communication subdomain compared to both other clusters. HC also showed a higher score in adaptive socialization and motor skills compared to MinR. All statistically significant results of analyses on the DQ and the VABS-II across the three subgroups at baseline are illustrated on Fig 2. There was no difference in the total ADOS CSS. In the ADOS subdomains, we found that HC exhibited lower RRB compared to MinR. Besides, the only difference between MinR and OptR was in global adaptive functioning (VABS ABC), MinR showing lower scores (70.7 ± 5.2) at baseline compared to OptR (78.5 ± 7.5).

**Figure 2:**
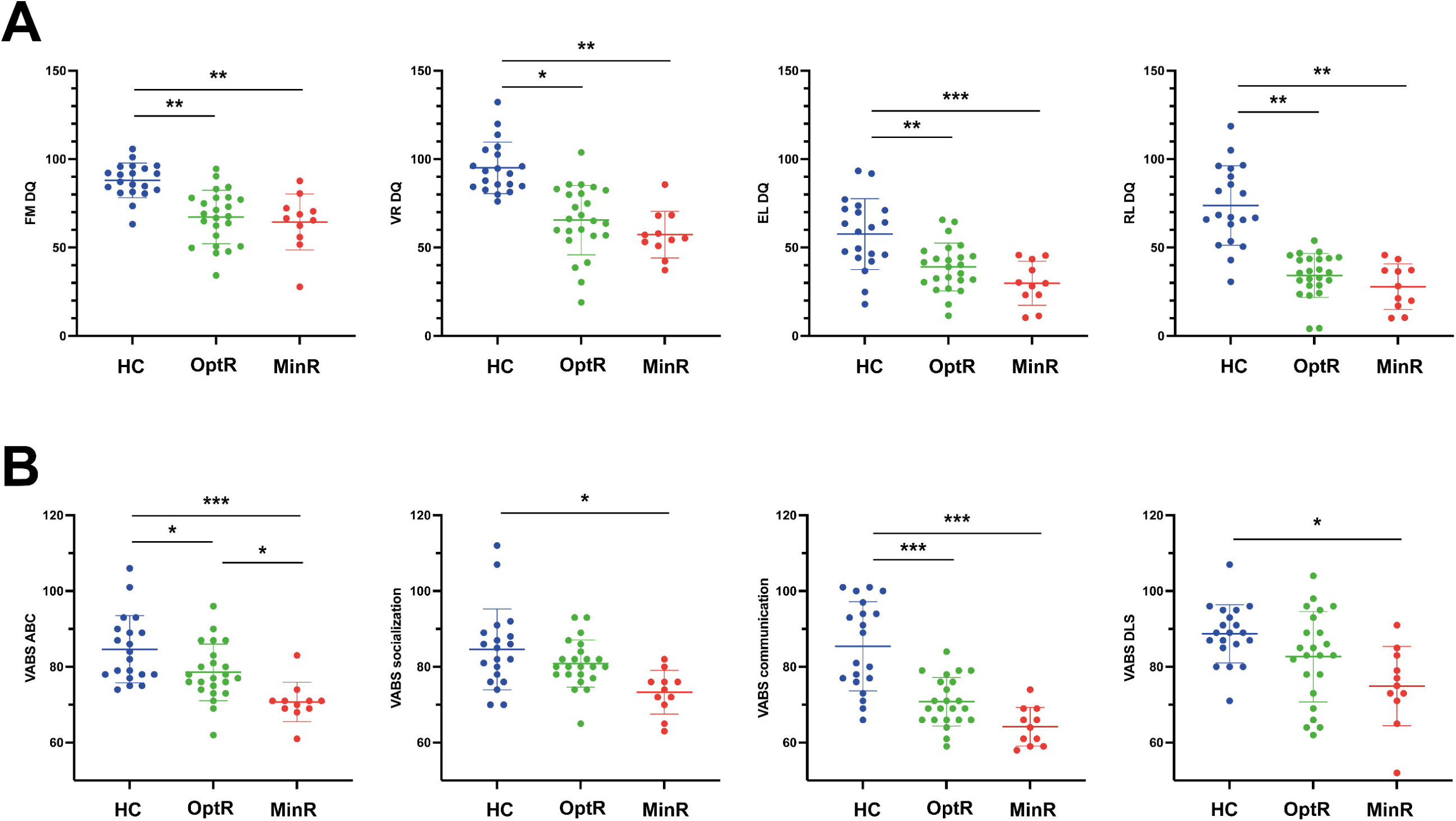
**A**. Statistically significant differences in DQ subdomains between subgroups at baseline. **B**. Statistically significant differences in VABS-II ABC and VABS-II subdomains between subgroups at baseline. \**p*<.05. \*\**p*<.01. \*\*\**p*<.001. ABC: Adaptive Behavior Composite. DQ: Developmental Quotient. EL: Expressive Language. FM: Fine Motor. HC: Higher Cognitive. MinR: Minimal Responders. OptR: Optimal Responders. RL: Receptive Language. VABS-II: Vineland Adaptive Behavior Scale. VR: Visual Reception.

#### Differences between subgroups in rates of change over 6, 12 and 24 months of intervention

We found that over the two years of therapy, OptR exhibited higher rates of change compared to the other two subgroups in cognition (composite DQ as well as VR, FM and RL subdomains). They also showed higher rates of change in adaptive behavior compared to MinR (VABS-II ABC, in socialization, communication and DLS subdomains) (see Supplementary table 2). These differences between MinR and OptR were already present within the 12 first months of therapy in the communication subdomain. Also, we found that MinR exhibited slower rates of change during the total time of intervention compared to both other subgroups in cognition (composite DQ, VR, FM and EL) as well as in adaptive behavior (VABS-II ABC, socialization, communication and DLS). Differences in the rates of change of composite DQ, VR and EL between MinR and OptR were already significant during the first year of intervention.

Finally, we found that OptR already exhibited faster rates of change in composite DQ, and adaptive functioning (VABS ABC) compared to MinR (Fig 3) after only 6 months of intervention. These differences in early DQ and VABS ABC rates of change were driven by RL and adaptive communication SPC.

**Figure 3:**
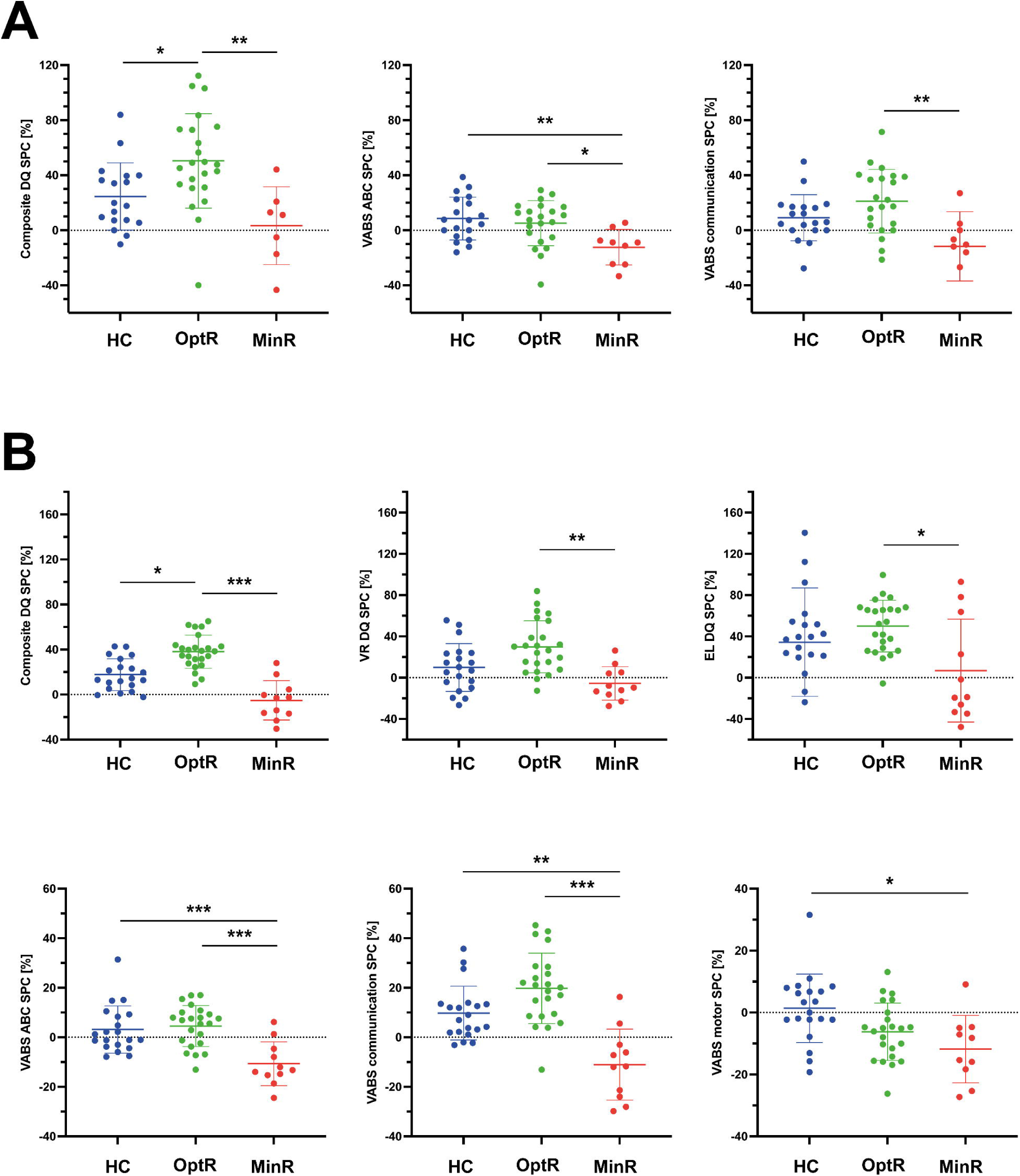
**A**. Statistically significant differences between subgroups in the rates of change of behavioral measures (DQ and VABS-II) within the first 6 months of intervention. **B**. Statistically significant differences between subgroups in the rates of change of behavioral measures (DQ and VABS-II) within the first 12 months of intervention. \**p*<.05. \*\**p*<.01. \*\*\**p*<.001. ABC: Adaptive Behavior Composite. DLS: Daily Living Skills. DQ: Developmental Quotient. EL: Expressive Language. FM: Fine Motor. HC: Higher Cognitive. MinR: Minimal Responders. OptR: Optimal Responders. RL: Receptive Language. SPC: Symmetrized Percent Change. VABS-II: Vineland Adaptive Behavior Scale. VR: Visual Reception.

We did not find any difference in the rates of change of symptom severity (total ADOS, SA and RRB) between the three subgroups during the time of intervention.

#### Subgroup classification based on early rates of change

Minimal responders (MinR) and optimal responders (OptR) showed no difference on the composite DQ at baseline and were both considered to have lower cognitive scores at baseline (LC). They were thus selected for our classification analyses to address the potential of clinical measures at baseline as well as their early rates of progress to classify them. At baseline, these two subgroups differed in VABS ABC. Logistic regression based on this parameter allowed an overall classification precision of 70.4% (5 out of the 11 MinR and 21 out of the 23 OptR were classified correctly). The model was significant (*χ*^*2*^ *= 10.0, p =* .*002*) and explained 35.5% of the variance (Nagelkerke *R*^*2*^).

Within the first 6 months of therapy, MinR showed slower SPC in the VABS-II ABC and in the composite DQ. Logistic regression based on these two variables allowed a partition of MinR and OptR with a 85.2% overall correct classification rate. Nineteen out of the 21 OptR included in the model and 4 out of the 6 MinR included were classified correctly. The logistic regression model was statistically significant (*χ*^*2*^ *= 10.2, p =* .*006*) and explained 48.0% of the variance (Nagelkerke *R*^*2*^). The prediction equation was the following: *0 = -0.040 * DQ SPC - 0.112 VABS-II ABC SPC -0.92*.

Within the first 12 months of therapy, OptR exhibited higher SPC in both the VABS-II ABC and the composite DQ. Logistic regression performed with both measures reached a 94.3 % rate of overall correct classification between OptR and MinR. Twenty-two out of the 23 OptR included in the model and 9 out of the 11 MinR included were classified correctly. The logistic regression model was statistically significant (*χ*^*2*^ *= 22.5, p < 0.001*) and explained 67.5% of the variance (Nagelkerke *R*^*2*^). The prediction equation was the following: *0 = -0.070 * DQ SPC - 0.167 VABS-II ABC SPC - 0.020*.

Ultimately, we combined the information at baseline with the rates of change to see if the classification model was enhanced (Figure 4). Combining VABS ABC at baseline with VABS ABC and DQ SPC within the first 6 months we achieved a model with 96.3% overall precision. Five of the 6 MinR and all of the 21 OptR were classified correctly (Nagelkerke *R*^*2*^ = 70.2% ; *χ*^*2*^ *= 16.6, p = 0.001* ; *0 = -0.268 * VABS-II ABC -0.049 * DQ SPC - 0.151 * VABS-II ABC SPC + 19.450)*. Combining VABS ABC at baseline with VABS ABC and DQ SPC within 12 months the model reached 94.1% overall precision. Ten of the 11 MinR and 22 of the 23 OptR were classified correctly (Nagelkerke *R*^*2*^ = 75.5% ; *χ*^*2*^ *= 26.4, p < 0.001* ; *0 = -0.188 * VABS-II ABC -0.040 * DQ SPC - 0.191 * VABS-II ABC SPC + 13.107)*.

**Figure 4:**
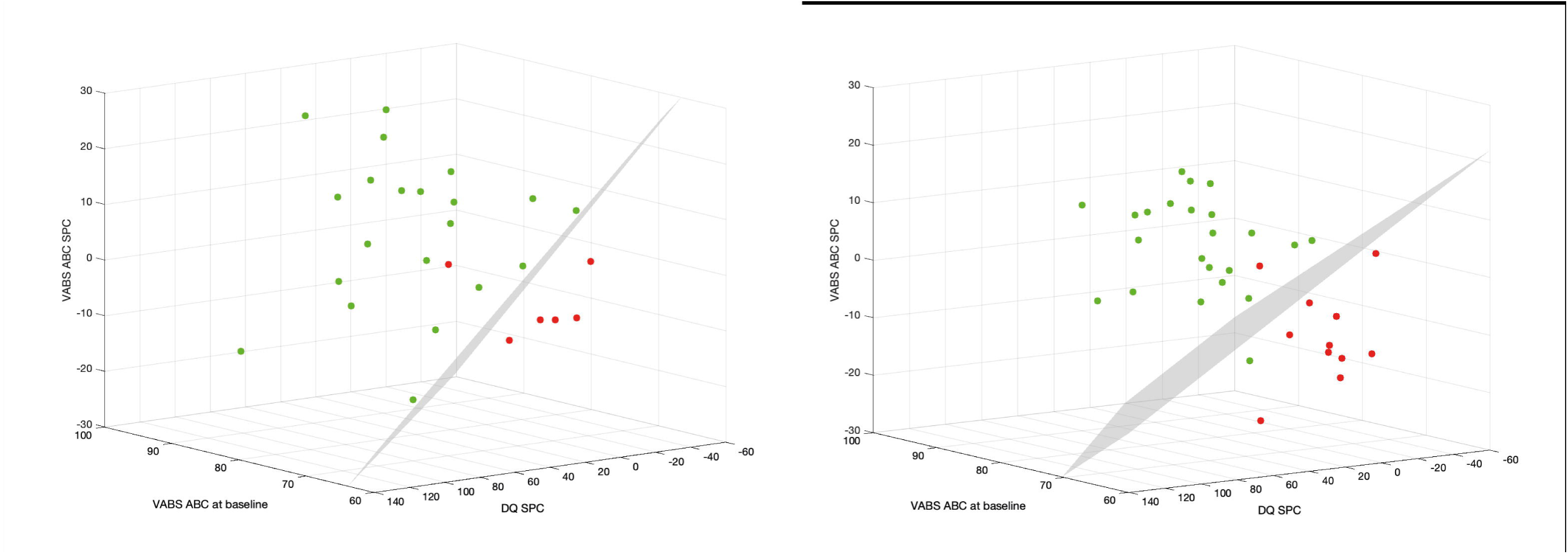
**A**. Multinomial logistic regression between MinR and OptR using their rates of change within the 6 first months of intervention in Composite DQ and VABS-II ABC, as well as their VABS-II ABC at baseline. Color code corresponds to subgroup membership (red: MinR, green: OptR). Decision boundary is represented in a dotted line. **B**. Multinomial logistic regression between MinR and OptR using their rates of change within the 12 first months of intervention in Composite DQ and VABS ABC, as well as their VABS-II ABC at baseline. ABC: Adaptive Behavior Composite. DQ: Developmental Quotient. SPC: Symmetrized Percent Change. VABS-II: Vineland Adaptive Behavior Scale.

In other words, it would have already been possible for a clinician to classify a child as being an OptR or MinR with 96.3% of accuracy after six months of intervention based on the child’s adaptive functioning at baseline and its rates of change in adaptive functioning and cognition. After 12 months of intervention and based on the same parameters, the overall precision classification would have reached 100%.

## Discussion

In the present study, we analyzed data from one of the largest samples of children who underwent two years of intensive (20h per week) and individualized ESDM intervention to identify predictors of their developmental outcome. Overall, we observed that, preschoolers in our sample made significant cognitive progress and adaptive skill gains over the two years of intervention (see Fig 1A). Improvements in the current sample allowed 72.7% of the children to enter a regular education classroom post-intervention, which in Geneva requires the child to have near peer-level functioning. These results are consistent with those reported in other studies on ESDM-based intervention (Dawson et al., 2010; SJ. Rogers et al., 2012). More specifically, our sample exhibited an average change in DQ (+19.9 points) that is very close to the one described in the randomized controlled trial (RCT) study by Dawson et al. (Dawson et al., 2010), which reported 18 points of cognitive gain, an average significantly greater than that of their control group. In parallel, a naturalistic study that explored developmental trajectories in preschoolers with ASD who were not enrolled in any specific therapeutic program reported an average DQ gain of only 6.3 points between 24 and 48 months of age (Clark et al., 2017). The average initial DQ in the cited study (63.6 ± 11.5) was similar to ours (60.3 ± 18.0). Considering similarities in the outcome between our results and previous ESDM studies as well as differences with naturalistic studies, one can infer that ESDM intervention in our study had a causal effect on the improvements observed at the whole group level. These results therefore highlight the possibility of implementing ESDM in Europe as effectively as in the US, despite differences in culture and health care system. They also support the cost-effectiveness of ESDM intervention, with improvements in cognition and adaptive behavior known to reduce subsequent school-based support needs, offsetting costs associated with early intensive intervention (Cidav et al., 2017; Penner et al., 2015; Peters-Scheffer et al., 2012).

This study also aimed to determine whether preschoolers with ASD who participated in a two-year NDBI intervention program (here ESDM in an individual setting, or I-ESDM) could be separated into distinct subgroups based on their cognitive trajectories over time. To achieve this, we used a *k*-means cluster analysis (CA) approach with cognitive abilities at baseline and cognitive rates of change over time as variables. CA yielded three groups: 36.4% (n = 20) of children with a mild cognitive delay at baseline that displayed a globally good outcome (Higher cognitive at baseline: HC), and two groups of children constituting the lower cognitive scores at baseline group (LC) that had very different outcomes. The first group of LC, which represented 43.6% (n = 24) of the entire sample, underwent significant cognitive and adaptive skill improvements (Optimal responders: OptR) while the second group of LC, which represented ∼20.0% (n = 11) of our sample, showed slower overall progress, and saw a widening of the developmental gap over time in cognition and adaptive behavior compared to same aged peers (Minimal responders: MinR). The clear distinction between toddlers with mild cognitive delay (HC) and those with the more severe cognitive delay at baseline (LC) observed in our sample is also reported in previous studies that applied CA to preschoolers with ASD (Fein et al., 1999; Stevens et al., 2000; Zheng et al., 2020). These studies identified at least two subgroups categorized as “high” and “low-functioning” based on early cross-sectional cognitive measures. One of the main differences in the present study is that we included a longitudinal variable in our CA (i.e., the rate of cognitive change) and were therefore able to further define our subgroups of LC children based on individual cognitive trajectories over time that a cross-sectional CA would have failed to capture. In our second analysis, we aimed to uncover potential predictors of outcome by evaluating how we can predict a child’s cluster membership. Amongst the LC subgroups (MinR and OptR), we found one difference at baseline in general adaptive functioning at baseline (see Supplementary Table 1). More importantly, we noted a significant difference between their rates of change in cognition and adaptive skills within the first 6 months of intervention. These differences were mostly driven by the progress in receptive language and adaptive communication. Using a logistic regression model, we showed that these early rates of change combined to differences at baseline predicted at 96.3 % attrition to either the MinR or OptR subgroups. This means that higher adaptive functioning skills at baseline combined to early, rapid developmental progress by 6 months of intervention allowed an accurate classification of subsequent developmental pattern.

Our analyses of the HC subgroup suggest that a mild cognitive delay (78.6 ± 10.9 of composite DQ) at the start of an ESDM intervention is associated with an alleviation of the delay in cognitive skills (+9.9% DQ per year) and adaptive behavior (+4.2% ABC per year) over the course of treatment. In addition, children in the HC group exhibited higher levels of adaptive skills compared to other subgroups (84.9 ± 9.2 of ABC) at baseline, especially in the VABS-II domain of communication (84.7 ± 8.9). All HC children except for one were able to continue into a regular education classroom following the intervention. With a DQ of 64 at both the beginning and the end of the intervention, this child was the only one in the HC group with a DQ value lower than 80 at the end of the intervention. Overall, our HC subgroup results suggest a positive outcome (in terms of cognition, adaptive functioning and schooling) in preschoolers with a mild delay in cognition and communication at baseline after receiving an individualized and intensive ESDM intervention. A recent review concluded that a higher cognitive level at baseline is a good predictor of positive outcome after various types of EI (Zachor & Ben-Itzchak, 2017). Also, previous studies focusing on another type of intervention (Applied Behavioral Analysis: ABA) reported that higher abilities in adaptive behavior (Eldevik et al., 2010; Sallows & Graupner, 2005) as well as in language (Magiati et al., 2011) constitute predictors of good outcome. This might suggest that mild delays in cognition and communication could represent a common predictor of good outcome among various EI approaches. These findings will need to be further explored with future RCT that assess the specific causality of ESDM intervention within these results. A practical implication of our findings concerning the HC subgroup is that clinicians who refer a toddler with a mild developmental delay at baseline to an ESDM program can be relatively confident that there will be a good outcome in cognition and adaptive behavior by the end of the intervention.

Apart from the HC group, the rest of the sample included children presenting a severe cognitive delay at baseline (Lower Cognitive, or LC). These children presented drastically different cognitive trajectories of change over time and were attributed to two distinct subgroups: OptR and MinR. Despite their severe cognitive delay at baseline (average DQ of 51.5 ± 10.7), the 24 children that composed the OptR subgroup greatly improved their cognitive and adaptive skills over time and 79.2% of them were able to join a regular education classroom with in-class support. On the other hand, the 11 children in the MinR subgroup had a similar level of cognitive impairment at baseline (average DQ of 44.9 ± 8.1), however their cognitive and adaptive functioning scores did not improve over time. Furthermore, the developmental gap between continued to widen, despite receiving intensive early intervention. Only 2 out of 11 (18.2%) MinR children joined a regular education classroom following the intervention. One clinical implication of our analyses of OptR and MinR at baseline is that the OptR constituted most of the LC children (68.6%) thus supporting the *a priori* that most toddlers who present with lower cognitive scores at intake display a positive outcome after receiving an individual intensive ESDM intervention. Nonetheless, a better understanding of the factors (behavioral, biological, and environmental) that are associated with MinR remains necessary to develop more targeted clinical recommendations. For instance, future studies including more participants with comorbid conditions such as epilepsy should investigate whether they represent moderator of outcome. Furthermore, they could determine whether the additive effect of various genetic mutations may moderate the outcome (Warrier et al., 2020). In our sample, four participants had a reported genetic finding with a potential causal effect in ASD. Two of them were in the MinR groum one in OptR and one in HC. Yet, the sample is far too small to draw any conclusion on the matter and future studies should address how genetic alterations modulate the RTT. The observation of two distinct trajectories of change in children with larger cognitive impairments at baseline could shed new light on the inconsistencies that exist between various studies that measured cognitive response to EI within LC preschoolers with ASD. For instance, one previous study concluded that children with this kind of profile only improve in fine motor skills and receptive language but not in adaptive behavior after receiving an early and intensive ABA intervention (Ben-Itzchak et al., 2014). Other studies focusing in ESDM reported an association between lower cognitive level at baseline and high cognitive gains (Devescovi et al., 2016; Robain et al., 2020). One can hypothesize that the inter-individual heterogeneity of outcome reported by previous studies (Ben-Itzchak et al., 2014; Devescovi et al., 2016; Robain et al., 2020), as well as the differences in their results were potentially due to the existence of two latent subgroups (MinR and OptR) that may have driven results in opposite directions. Our results thus advocate for a more systematic subgroup phenotyping, including longitudinal variables, in future studies focusing on the clinical outcome of EI to better describe the phenotypic heterogeneity within LC preschoolers with ASD.

Finally, our results suggest that the outcome after two years of intervention for children with LC at baseline can be predicted by the end of the six first months of intervention with high accuracy. Adaptive functioning was the only clinical parameter that could help distinguish OptR from MinR at baseline, allowing an overall classification precision of 70.4%. Nonetheless, based on this single variable only 45.5% of MinR could be classified correctly resulting in a relatively poor sensitivity in MinR identification at baseline. Sensitivity to OptR was largely higher with 91.3% of them correctly classified at baseline. Thus, the clinical interest for using the VABS-II alone at baseline to discrimate between LC (OptR and MinR) appears very limited. Nonetheless, the OptR group’s rates of change appear to be significantly higher than those observed in the MinR group within the first year for cognitive and adaptive skills (especially in communication). Our results are in line with those of Sallows et al. (Sallows & Graupner, 2005), who reported cognitive gain during the first year of an early and intensive ABA intervention as one of the best predictor of outcome at the end of the intervention. Furthermore, we found that based on adaptive functioning at baseline combined to the rates of change in cognition and adaptive functioning within the first six months of intervention, we could infer the outcome after two years of intervention. Together, these conclusions might shed light on the timing of RTT, and when children can be considered as “*non responders*’’ as raised by Vivanti et al. (Vivanti et al., 2014). Indeed, our analyses suggest that the first 6 months to a year of intervention offers critical information about how a child will respond to ESDM intervention over time, and leads us to question whether an early, clear response to intervention can predict an optimal response overall. The emphasis on this early response to intervention as a predictor of long-term outcome has several clinical implications. One of them would be the importance of implementing regular, standardized follow-ups to measure children’s cognition and adaptive behavior in the first 6 and 12 months, in addition to the systematic ESDM *Curriculum Checklist* (ESDM-CC) that is currently used in the model. An alternative could lie in the development of a standardized way to use the ESDM-CC to track the rate of developmental change and ultimately the post-intervention outcome. This type of early standardized follow-up could potentially alert the clinician of difficulties a MinR child might face. This does not mean that MinR should be given less resources in terms of intervention. Our results show that despite a less optimal response compared to others, MinR show improvements in their raw scores. In the perspective of personalized medicine, future studies should determine what is the best intervention for these children. It might be possible that they would benefit from earlier or longer or more intensive intervention. It could also be that another type of EI (other than ESDM) would provide them a more optimal outcome. More research is needed to understand what supports or program enhancements would allow a child with a slower response to intervention to have a more optimal outcome.

In summary, our results show that despite the lack of individual reliable predictors of outcome for children with ASD who present severe cognitive delays at baseline, the consideration of their early dynamic behavioral parameters may help predict their overall response to intervention. Further RCTs that explore the trajectories of subgroups similar to ours are needed to determine the precise effect of the ESDM on children with MinR and OptR profiles. More specifically, we need to understand whether ESDM helps OptR improve their outcome or if it prevents MinR from falling even further behind developmentally, or both. Another hypothesis to be addressed is whether ESDM has an influence in the relative number of participants that are affected to each subgroup – i.e., whether some OptR participants would have been MinR if they had not undergone an ESDM based therapy. Future research on the specific effects of ESDM on each subgroup could result in improved therapeutic guidelines that are more tailored to each child’s individual developmental trajectory. Our study provides relevant variables that should be explored by future research at the beginning and during the very first months of an ESDM intervention.

### Limitations

Despite being one of the largest samples of preschoolers who benefited from a two-year intensive and individualized ESDM program, the sample size of the present study limits the number as well as the size of subgroups that can be detected by a cluster analysis. Nevertheless, we took care to respect the commonly accepted prerequisites of cluster analyses, including the minimum sample size in each group or the number of factors in the analyses given the overall sample size (Formann, 1984; Qiu & Joe, 2006). It is possible that studies performed on larger samples could achieve more fine-grained subgrouping on a similar population based on the same measures and could lead to bigger subgroups, in turn increasing the statistical power to detect differences at baseline between lower cognitive clusters that we could not highlight.

Another limitation that is a direct consequence of the previous one lies in the choice of the main outcome. We chose parameters related to cognitive skills as the main clustering factors. However, it would have been possible to use other measures such as level of ASD symptoms, adaptive skills or even a combination of these two. The inclusion of more variables in the model could help in defining a larger number of clusters and therefore increase our understanding of the heterogeneity of ASD in a refined manner. However, this was not possible in the present study, because of the limited sample size. The addition of more variables in the model and the multiplication of clusters would have violated the cluster analysis assumptions, making its interpretation invalid. Studies with larger samples should include more clinical parameters and could also use outcome variables suggested by parents (McConachie et al., 2018).

Within our sample, 7 children did not have their DQ at baseline assessed with the same test as the rest of the sample. Indeed, these 7 children were tested with the PEP-3 while the others were tested using the MSEL. Although the scores obtained via these two assessments show a strong consistency within our sample (Cronbach α = .914, n=44), it is not possible to affirm that they are equivalent due to their different design. Yet, the clustering analysis applied on the sample with the 7 children excluded yielded the same cluster solution. Nonetheless, this divergence in the test used for a minority of our children should be kept in mind when interpreting the results.

A last limitation here lies in a lack of a non-ASD lower cognitive (LC) control group making difficult to evaluate the causality of ESDM intervention in the observed outcome of this specific population. Nevertheless, Hedval et al. reported that 87.7% of the preschoolers with ASD and LC at baseline (<70 of DQ) still had a DQ lower than 70 when assessed after two years without receiving any EI (Hedvall et al., 2014). Moreover, their delay in adaptive functioning worsened in all the VABS-II subdomains except for communication at the group level. In contrast, in the present study LC children with similar developmental pattern (MinR) only constituted 31.4% of our LC group, while the other LC participants (OptR) exhibited large improvements in DQ as well as in adaptive behaviors. Considering these results, one can infer a causal effect of ESDM in the progress made by children with important cognitive delay at start. The specific effects of ESDM compared to other types of EI still needs to be addressed with future RCT.

## Conclusion

In this study, we applied a cluster analysis to the largest European sample of preschoolers with ASD who participated in an ESDM program for 20 hours a week over a two-year period. Overall, we found that ASD symptom severity decreased, and cognitive delay improved over the intervention period. Furthermore, the cluster analysis suggested three main patterns of cognitive trajectories over time. First, children who displayed mild cognitive and adaptive behavioral delays at baseline tended to have a good developmental prognosis, finishing their two years of early intervention with cognitive and adaptive behavior scores within the normal range. Second, children who presented with severe cognitive delays at the start of their early intervention exhibited two dramatically different patterns of developmental trajectories. About a third of these children continued to fall behind developmentally, despite intensive therapy services. The two remaining thirds of the children, who presented with lower cognitive and adaptive behavior scores at the beginning of treatment, exhibited early and important gains in cognition and adaptive behavior which continued for the duration of the 2 years of intervention. We found that the two lower cognitive subgroups differed in their global adaptive functioning at baseline, although this parameter alone shows a limited sensitivity in identifying the children who will show slower gains. Nevertheless, our results suggest that it may be possible to predict, after only 6 months of early intervention, and with very high levels of accuracy, whether a child will have an overall minimal or optimal response to treatment, based on their early gains in cognition and adaptive behavior combined to their adaptive functioning at baseline. These results advocate for close monitoring using standardized cognitive and adaptive behavioral testing during the first six months of intervention, especially for children that exhibit a clinically significant cognitive delay at baseline. Having an understanding early-on of how a child is responding to early intervention could alert clinicians and parents to the need to adapt and enhance the child’s treatment plan. Future studies are needed to replicate these findings, and to evaluate the kinds of treatment adaptations that would optimize child outcome for each ASD subgroup. Also, there is a need for longitudinal studies that provide a long-term follow-up in the years following the end of early intervention, to be able to assess whether the patterns of cognitive profiles and response to treatment observed remain stable over time. Overall, our results advocate for a more systematic use of subgroup phenotyping that includes longitudinal parameters when assessing the efficacy of an early intensive intervention, to better decipher the great heterogeneity of behavioral dynamics in treatment response.

## Data Availability

The datasets generated in the current study are available from the corresponding author on reasonable request.

## Supplementary Material

Supplementary table 1

Supplementary table 2

Supplementary table 3

## Declaration

### Author’s contribution

M.S. conceived and designed the study. M.F, M.G., N.K., and F.R. participated in the data acquisition. M.G. and F.R. prepared and analyzed the data under the supervision of M.S. All authors participated in interpretation of results. M.G. and F.R. wrote the manuscript with the inputs from all other authors. All authors read and approved the final manuscript.

## Acknowledgments

The authors would like to thank all the families who kindly participated in the study, all therapists at the *Centre d’Intervention Précoce en Autisme* in Geneva, as well as Alexandra Bastos, Stéphanie Baudoux, Lylia Ben Hadid, Aurélie Bochet, Léa Chambaz, Flore Couty, Sophie Diakonoff, Lisa Esposito, Constance Ferrat, Marie-Agnès Graf, Oriane Grosvernier, Kenza Latrèche, Sara Maglio, Matthieu Mansion, Eva Micol, Irène Pittet, Sonia Richetin, Laura Sallin, Stefania Solazzo, Myriam Speller, Chiara Usuelli and Ornella Vico Begara for their help with data collection.

## Ethics approval and consent to participate

Informed consent was obtained from the parents of all participants included in the study. Swissethics - Commission d’éthique Suisse relative à la recherche sur l’être humain approved this study (Protocole 12-163/Psy 12-014), referred under the number PB_2016-01880 and accepted on september the 25th, 2012.

